# Retrospective Study of an Epidemic *Vibrio Cholerae* in the Central Region of Ghana; An Evidence from Surveillance Data

**DOI:** 10.1101/2023.07.22.23293033

**Authors:** Gideon K. Acheampong, Isaac Owusu, Fidelis Zumah, Ernest Akyereko, Rebecca Ann Mpangah

## Abstract

**Background:** In the Central region of Ghana, cases of cholera were detected in October 2016, in the Cape Coast Metropolis. The number of cases detected in the peri-urban communities rose exponentially indicating a high transmission potential of infections. We conducted a descriptive analysis of surveillance data of the 2016 cholera outbreak in the Central Region with the aim of describing the epidemiological features of the outbreak.

**Methods:** A retrospective analysis of cholera cases between October and December 2016 was conducted using variables including date of onset of symptoms, age, sex, rapid diagnostic test (RDT) results and district of residence of cases. Cases were descriptively characterized in terms of time, place, and person, attack rates were computed, and an epidemic curve was constructed using the date of onset of symptoms of cases. Pearson’s chi-square/Fisher’s exact tests were used to determine associations among selected variables of cases.

**Results:** A total of 731 cases of cholera were reported with an overall attack rate of 67 cases per 100,000 population; no fatalities were recorded. The epi-curve showed multiple progressive peaks denoting a propagated type of outbreak driven by person-to-person transmission of infections. The mean age was 23 years, with 40% of cases occurring in the age group 15-24 years. The difference between the number of cases for males and females was not significant *(p-value = 0.619*). Close to 90% of all cases were reported from the Cape Coast Metropolis. Abura-Asebu-Kwamankese (AAK) and Komenda-Edina-Eguafo Abirem (KEEA) had a combined number of 64 cases (10%). There was a significant association between RDT results and the bacterial culture test (p<0.001), as well as that between sex and final case classification *(p=0.004)*.

**Conclusion:** The cholera outbreak affected a total of 731 people, with the highest number of cases reported in the 15-24 year age group. The outbreak was driven by person-to-person transmission and contaminated food and water sources. Rampant open defecation, open roadside food and water vending, and poor personal hygiene practices including hand washing were identified as major risk factors. The Cape Coast Metropolis and the KEEA were the most affected with the highest number of cases and the highest attack rate. The outbreak was predominantly confirmed through rapid diagnostic tests and culture confirmation. Current and future development projects must be geared towards effective town planning and decongestion, provision of designated dumping sites, toilet facilities and more water treatment plants. It is also imperative that district health officials also explore the issues of poor health-seeking behavior and access to care as possible factors contributing to high morbidities.

## INTRODUCTION

Cholera is an acute intestinal disease caused by the bacterium *Vibro cholerae*. The disease is characterized by severe watery diarrhea with vomiting and dehydration. It is spread through fecal-oral route from an infected person. [1]–[3]. Cholera epidemics have high potential for spreading within the shortest possible time. Deaths typically result from dehydration that accompanies the acute severe diarrhea, which can be successfully treated with timely administration of oral rehydration salts (ORS). Cholera continues to be a global health threat. Pandemics of cholera have been experienced in some countries with sporadic attacks throughout the world, especially in areas where water supply, sanitation, food hygiene and safety continue to be a challenge. It affects all ages and both sexes with attack rates usually highest in children living in cholera-endemic areas. In 2014, there were 58 public health events within the World Health Organization (WHO) African Region and out of these, infectious diseases formed 95% of all these events with Cholera being the most frequently reported (31%), ahead of Ebola (13%) which has seen its biggest epidemic in history recently [4]. According to the WHO in 2015, new major outbreaks of cholera are continuing to occur, especially in the wake of climate changes. There were 105,287 cholera cases of which 1,882 resulted in deaths giving rise to a Case Fatality Rate (CFR) of 1.8% within the African Sub region [4]. These reported cases were more than double that of the previous years 2013 and 2014. In total, 16 countries reported cholera cases of which Ghana was the second most affected country with 28,944 reported cases with CFR of 0.8% only topped by Nigeria with 45,996 cases representing 2.1% CFR while DR Congo had 22,203 reported cases with 1.7% CFR. Ghana, Nigeria and DR Congo accounted for 85% of all cases reported in 2014 [4]. Cholera became endemic in parts of Ghana and the country experienced outbreaks of the disease every five years from 1970 to 2016 [5]. In June 2014, the country reported 6 cases of the disease in the Greater Accra region after 23 weeks of no confirmed case in the year. Within two weeks the number of reported cases had risen above 250 and began spreading to other regions [1], [6], [7]. By the close of 2014, a cumulative total of 28,955 cases with 243 deaths and a CFR of 0.8% were recorded. All ten regions in Ghana reported cases, with 70% of all the cases from the coastal areas alone (Central and Greater Accra Regions).

In the Central region of Ghana, a patient presented on 17^th^ October, 2016 to XX with acute watery diarrhea that was confirmed as cholera on XX. Over the next 5 days, two additional patients, also from Cape Coast Metropolis, presented with similar symptoms, that were subsequently confirmedvto be cholera. The number of cases detected in the peri-urban communities in Cape Coast Metropolis rose from 3 new cases recorded on 24^th^ October to 34 new cases identified on 26^th^ October, 2016. The exponential increase in cases indicated a high transmission potential of infection in the peri-urban communities. We conducted a descriptive analysis of surveillance data of the 2016 cholera outbreak in Cape Coast with the aim of describing the epidemiological features of the outbreak. Specifically, we set out to characterize the incidence by date of disease onset; describe the age and sex distribution of cholera cases; provide an overview of the distribution of cases across affected districts. A critical study of these issues is essential to provide information for future response to cholera outbreaks.

## STUDY SITE

The Central region occupies an area of 9,826 square kilometers or 4.1 percent of Ghana’s land area, making it the third smallest in the region after Greater Accra and Upper East regions. It shares common boundaries with the Western Region on the west, Ashanti and Eastern Regions on the north, and Greater Accra Region on the east. On the south is the 168-kilometre length Atlantic Ocean (Gulf of Guinea) coastline [8].

**Figure 1.**
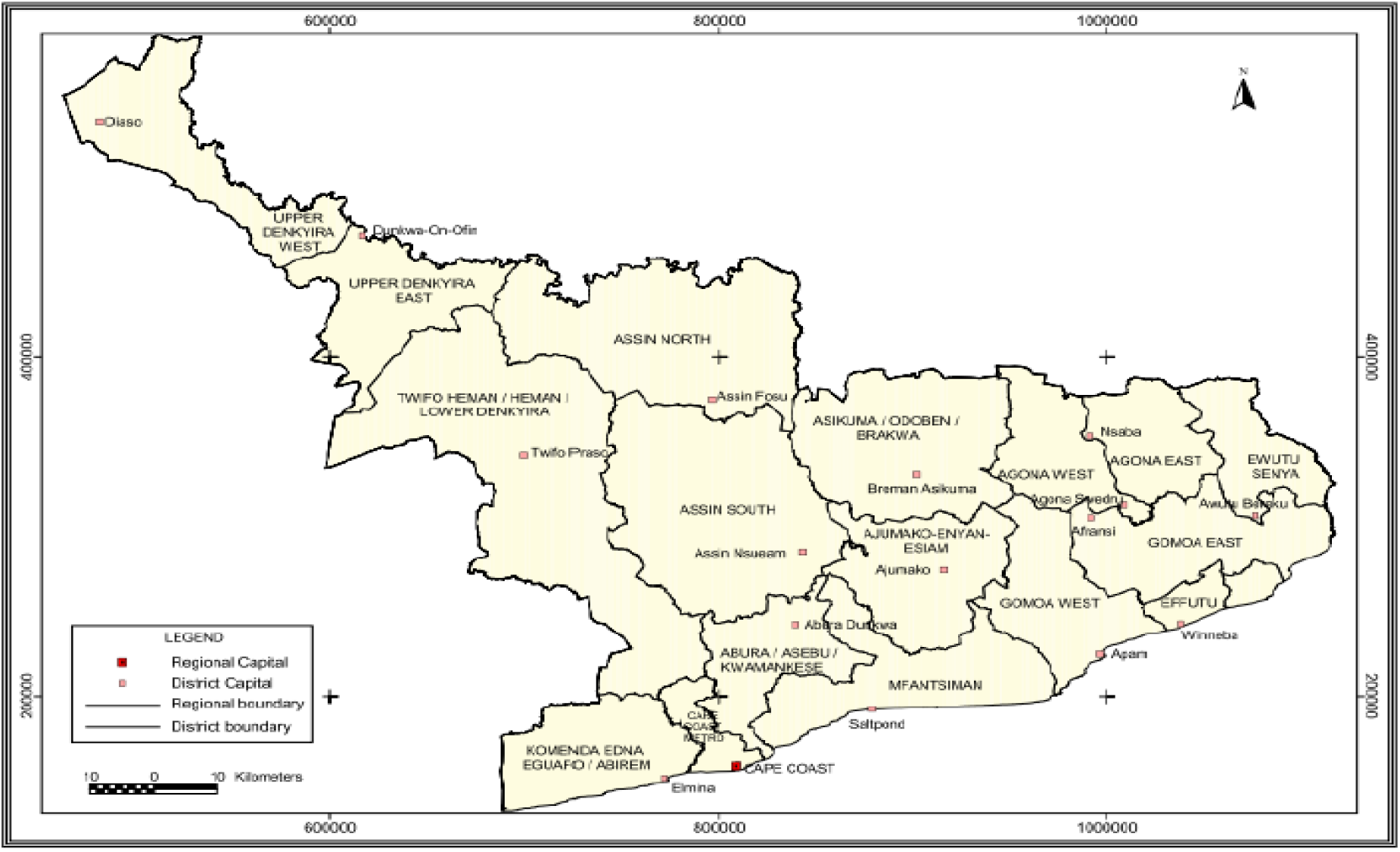
Map of the Central Region of Ghana.

## METHODS

A retrospective analysis of cholera cases from selected districts between 1^st^ October and 31^st^ December, 2016 from the Central Region was conducted using variables including date of onset of symptoms, age, sex, rapid diagnostic test results, final case classification (suspected, probable, confirmed), district of residence of cases, and bacterial culture results. The line list of all cholera cases recorded in the affected districts were compiled by the Disease Surveillance Department of the Ghana Health Service.

All data was double entered into Microsoft excel. The districts were namely, Abura Asebu-Kwamankese (AAK), Asikuma-Odoben Brakwa (AOB), Cape Coast Metropolis, Komenda-Edina-Eguafo-Abirem (KEEA), Mfantseman, Twifo-Hemang-Lower Denkyira (THLD) and Twifo Atti Mokwa (TAM). *Based on the definition provided in the National Technical Guidelines of the Integrated Diseases Surveillance and Response (IDSR), the following were established. A suspected cholera case was defined as any patient aged 2 years or older presenting with acute watery diarrhea, with or without vomiting, during the period of the outbreak. A confirmed case was defined as a suspected case with vibrio 01 or 0139 confirmed by culture*.

Stata statistical software package (version 12, StataCorp LP, College Station, TX, USA) was used for all univariate and bivariate analysis. Cases were descriptively characterized in terms of time, place, and person. An epidemic curve was constructed using the date of onset of cases over time. All cases with incomplete information were excluded from epidemiological analysis. All cases with incomplete information were excluded from the determination of attack rates and epidemiological analysis.

## RESULTS

A total of 731 cases of cholera were reported between 10^th^ October to 21^st^ December,2016 in the Central Region **[Figure 2]**, out of which 673 cases had data on age, sex and district of residence with 670 cases having information documented evidence on the date of onset of symptoms for cholera disease. Out of the 673 cases, 208 (31%) occurred in the age group of 15-24 years, and this was the predominant age group affected by the outbreak **[Table 1]**. Cases above 60 years formed 7% of the 673 cases while 9% of cases were children under 5 years of age. The mean age of cases was 23 years (range: 1 to 77 years) with a greater number of the cases being male (53%).

**Table 1.**
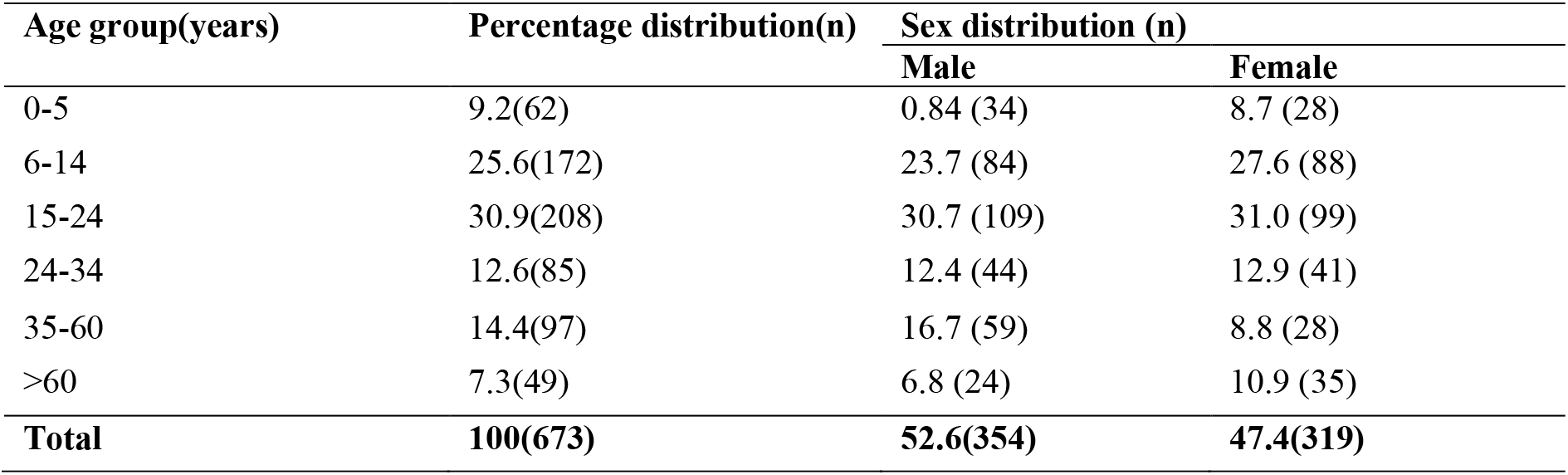
Host age and sex distribution of cholera cases recorded in the Central Region.

**Figure 2.**
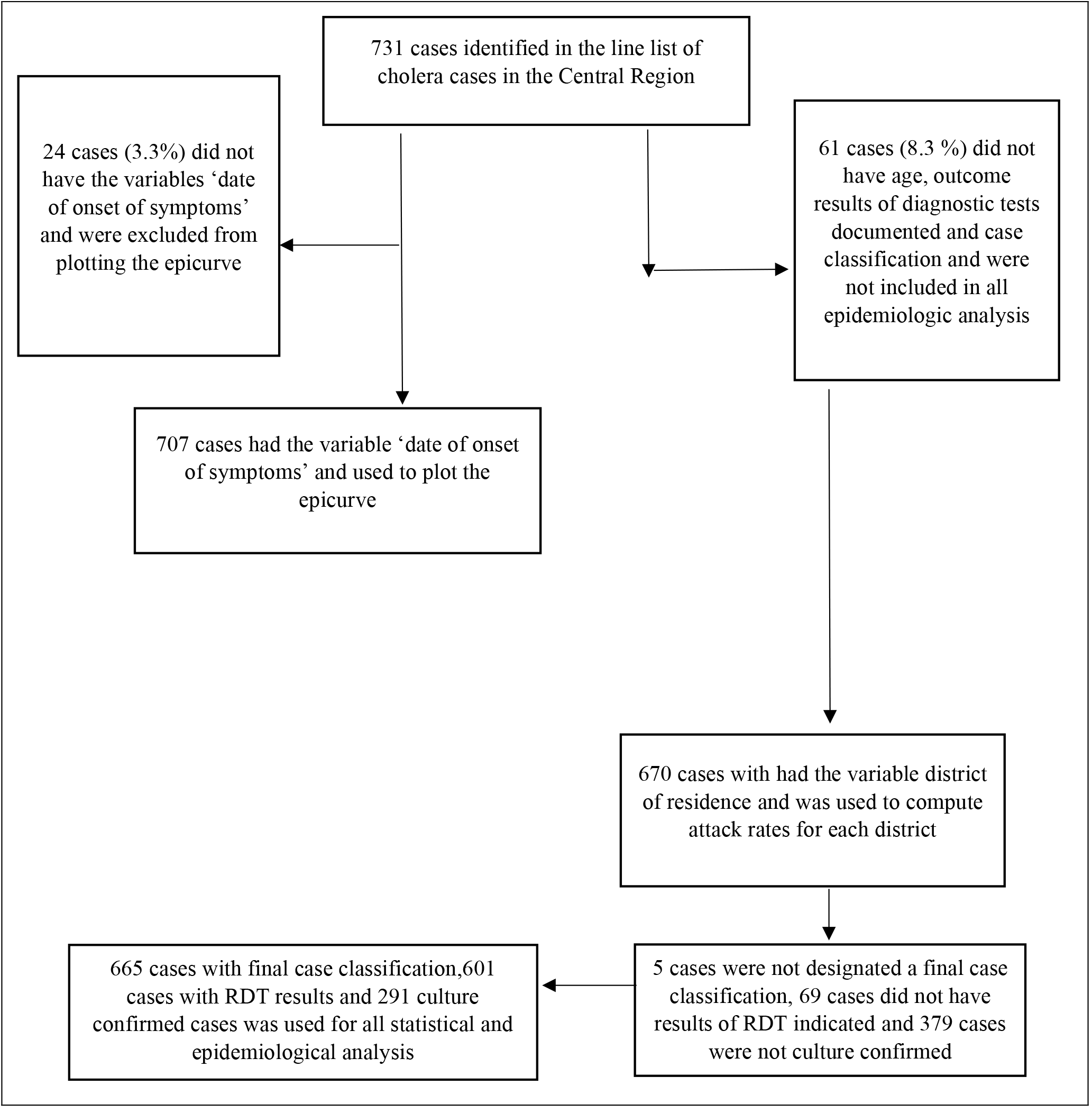
Flow chart of cholera cases reported in the Central Region during the outbreak.

**[Table 2]**. A total of 665 cases were classified based on either RDT results or culture confirmation, 601 cases with RDT results and 291 cases culture confirmed [**Figure 2]**. Out of the 665 cases that were classified, 350 of these cases were classified as suspected cases, 209 were classified as probable cases with only 106 confirmed as cases of cholera, this displayed in Figure 2 and Table 3. From Table 3, 285 cases out of 601 cases tested positive after a rapid diagnostic test was conducted, 118 of cases were culture confirmed to be cholera cases out of 209 subjected to bacterial culture test. Out of 670 cases with districts of residence indicated, 600 cases, representing 90% of all cases were reported from the Cape Coast Metropolis, the district with the highest number of cases. Abura Asebu-Kwamankese and Komenda-Edina-Eguafo-Abirem had a combined number of 64 cases, representing 10% of all reported cases with documented district of residence. An overall attack rate of 67 cases per 100,000 population was obtained as displayed in Table 4, the Cape Coast Metropolis was the district with the highest attack rate (285 cases per 100,000 population) followed by Komenda-Edina-Eguafo-Abirem (23 cases per 100,000 population) and Abura Asebu-Kwamankese (15 cases per 100,000 population).

**Table 2.** Summary age description of all cholera cases recorded.

| Variable | Min | Max | Mean | SD | Variance | Range | IQR |
| --- | --- | --- | --- | --- | --- | --- | --- |
| Age | 1 year | 77 years | 23 years | 18.03 | 325.29 | 76 | 19.5 |

**Table 3.** Frequency distribution of cases of cholera by districts of residence, test results and case classification.

| District of residence | Rapid diagnostic test results |  | Culture results |  | Final case classification |  |  |
| --- | --- | --- | --- | --- | --- | --- | --- |
|  | Positive | Negative | Positive | Negative | Suspected | Probable | Confirmed |
| ABURA ASEBU-KWAMANKESE | 4 | 12 | 2 | 2 | 15 | 5 | 2 |
| ASIKUMA-ODOBEN BRAKWA | 0 | 1 | 0 | 1 | 1 | 0 | 0 |
| CAPE COAST METROPOLIS | 267 | 275 | 110 | 152 | 302 | 198 | 100 |
| KOMENDA-EDINA-EGUAFO-ABIREM | 9 | 28 | 6 | 18 | 32 | 6 | 4 |
| MFANSTEMAN | 2 | 0 | - | - | 0 | 2 | 0 |
| TWIFU-HEMANG-LOWER DENKYIRA | 2 | 0 | - | - | 0 | 2 | 0 |
| TWIFO ATTI MOKWA | 1 | 0 | - | - | 0 | 1 | 0 |
| **TOTAL** | **285** | **316** | **118** | **173** | **350** | **209** | **106** |

**Table 4.** Attack rates stratified by districts.

| DISTRICT | CASES | PROJECTED 2017 POPULATION | ATTACK RATE/100,000 POPULATION |
| --- | --- | --- | --- |
| ABURA ASEBU-KWAMANKESE | 22 | 145,112 | 15.2 |
| ASIKUMA-ODOBEN BRAKWA | 1 | 139,566 | 0.7 |
| CAPE COAST METROPOLIS | 600 | 210,382 | 285.2 |
| KOMENDA-EDINA-EGUAFO-ABIREM | 42 | 179,190 | 23.4 |
| MFANSTEMAN | 2 | 178,729 | 1.1 |
| TWIFU-HEMANG-LOWER DENKYIRA | 2 | 68,270 | 2.9 |
| TWIFO ATTI MOKWA | 1 | 76,457 | 1.3 |
| **TOTAL** | **670** | **997,706** | **67.15** |

The epi-curve in Figure 3 shows multiple progressive peaks denoting a propagated type of outbreak driven by person-to-person transmission of infections, exacerbated by episodes of common-source infections such as contamination of food and/or water. The first peak was observed on October 26^th^ followed by the 2^nd^,3^rd^ and 4^th^ on November 11^th^,17^th^ and 27^th^ respectively. The final peak was then observed on the 16^th^ of December followed by a gradual decline in the number of cases with no new cases recorded on 21^st^ December,2016. The exposure factors included rampant open defecation, open roadside food and water vending, and poor personal hygiene practices including hand washing. These factors appeared to be the major drivers of the cholera outbreak in Cape Coast.

**Figure 3.**
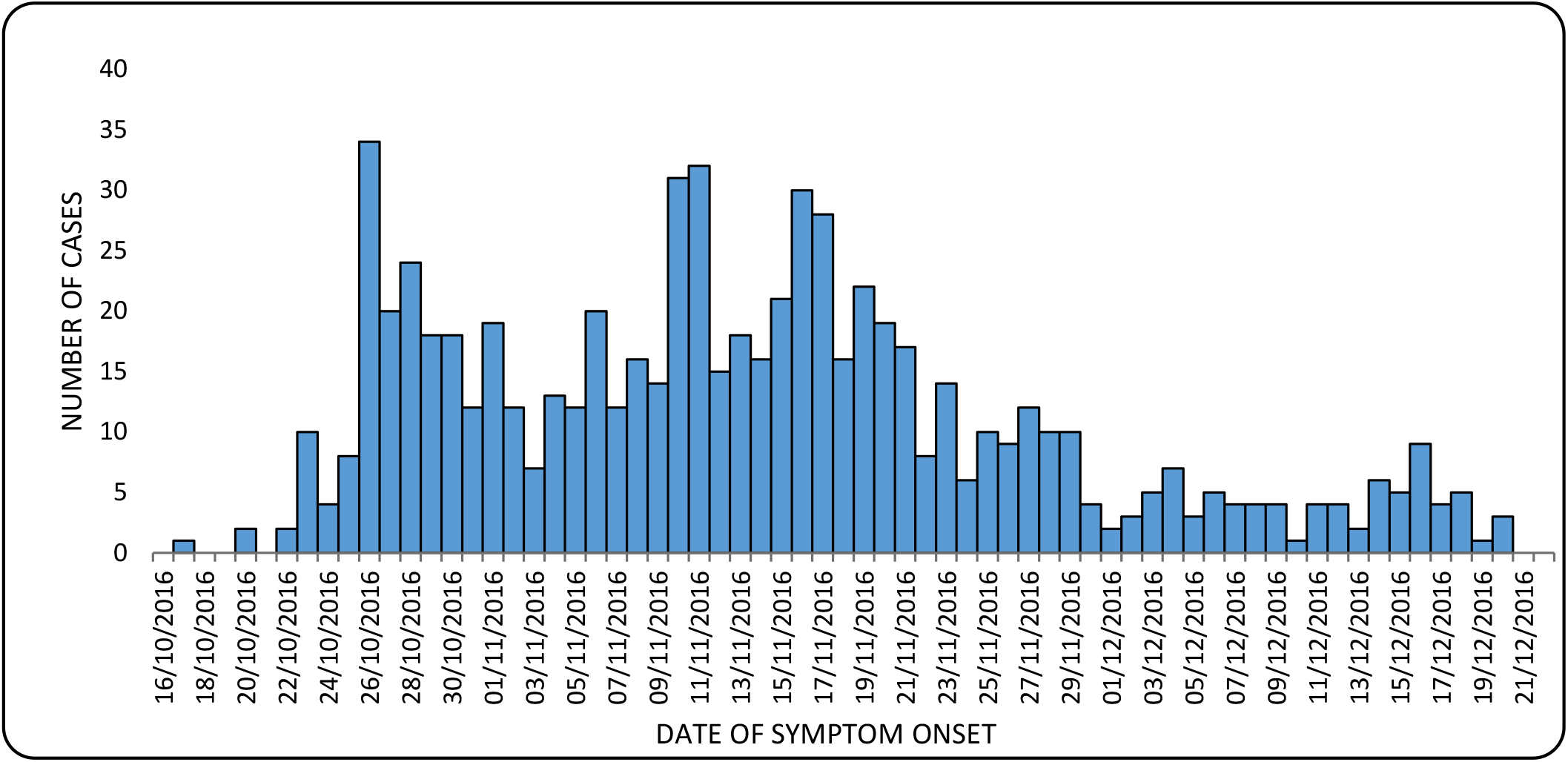
Daily number of reported cases by date of onset of cholera disease.

## DISCUSSION

Cholera is a major public health concern because of its high transmissibility, death-to-case ratio and ability to occur in epidemic forms[9], it remains a concern in many parts of Ghana especially in the coastal sectors[10] as well as among many poor and unsanitary communities, despite the detailed understanding of the bacteriology, epidemiology, and public health aspects for more than a century[11]. The currents descriptive study analyzed a large cholera outbreak in the last quarter of 2016 in the Central Region of Ghana, this was based on data obtained from 7 affected districts in the Region in subject. As previously mentioned, this was a propagated type of outbreak driven by person-to-person transmission of infections, escalated up by episodes of common-source infections such as contamination of food and/or water. A grand total of 731 cases were identified with no fatalities recorded.

High transmissibility and attack rates as in the case of this outbreak have been associated with limited access to health care, insufficiencies of the health care system and limitations in the surveillance system capacities to trigger timely response [12]–[14]. With proper and timely case management and more importantly a more efficient surveillance system cholera transmissibility can be reduced and any likely fatalities prevented. The results from the study also revealed that rampant open defecation, open roadside food and water vending, and poor personal hygiene practices including hand washing were common among the residents of the Central Region and served as risk factors for cholera disease. These unsanitary practices affect individuals’ hygiene and is as well a cause of water source contamination. Individuals from households without toilet facilities were more likely to develop cholera or diarrhoea compared with individuals from households with latrine facilities. The presence of toilets increases the chance of safe disposal of feces, one way to decrease contact between the causative organisms of cholera and the host [15]. In cases where children play in the dirt which may be contaminated by fecal matter, the dirt act as a reservoir of pathogens [15]–[17].

A further issue that could have contributed to the high transmissibility and morbidity of the outbreak was the unavailability of treated water increasing the likelihood of unsafe drinking water. The Central Region, one of the more rural regions in Ghana, is one of the more problematic regions in terms of water-related diseases due to its rural nature [18]. Current and future development projects must address the lack of sanitation through more water treatment plants and the use of rainwater harvesting systems. It is however imperative to note that comprehensive diarrheal disease prevention is not easy in poor communities, changing beliefs and habits, such as avoiding open defecation is difficult when there are no latrines close to markets, farms and other areas of work. In response to this development, social marketers should design interventions to target individuals with no formal education by educating them to change their behavior. In addition, social marketers should incorporate interventions aimed at enhancing individual’s self-efficacy. They should also raise awareness on perceived susceptibility, perceived severity, perceived benefits and cues of action. These will motivate individuals to engage in a healthier life style and change behavior towards cholera prevention [19]. It is also imperative that district health officials also explore the issues of poor health-seeking behavior and access to care as possible factors contributing to high morbidities and deaths in the community. It is key to highlight the importance of providing regular community education on cholera signs and symptoms and prevention and treatment measures even in periods outside epidemics [10].

With regards to sex distribution of cases, more males were affected by females, it is however worth noting that the difference between the number of cases for males and females is not entirely significant, a case which is further buttressed by the p-value of the Fisher’s exact test for association between sex and the results of the bacterial culture test of cases (p-value = 0.619). However, a likely explanation for the high number of males affected is the fact that males were more likely to purchase food from road side vendors, a group that has been identified as exposure factors for the spread of the outbreak [11]. On the other hand, females traditionally are more involved in fetching water for domestic use from any likely local stream, possibly drinking directly from the stream, which could expose them further to the infection [11]. A number of findings have documented that children below the age of 5 years have highest incidence of cholera, age-specific mortality is as well highest in this age group [20]–[22]. In this outbreak however, a different pattern was observed, majority of cases were recorded in age groups 15-24 years (31%) followed by the 6-14 years (26%) age category. A mean age of 23 years indicates that majority of affected persons were adults in that they had increase exposure to *V. cholerae*.

In a further development, some studies have demonstrated that some cholera outbreaks have been caused by multidrug resistant atypical *V. cholerae* O1 and EI Tor strains, which are reportedly highly virulent [23], [24]. Vibrio cholerae El Tor O1 Ogawa was responsible for the endemics in Nepal before 2012 and previous outbreaks of cholera in Kathmandu valley in 2004 [23]. Proper antibiotic susceptibility testing of V. *cholerae* is important to guide appropriate antimicrobial therapy. Guidelines for managing and containing cholera outbreaks which include, in addition to rehydration, using the antimicrobials trimethoprim, and more recently ciprofloxacin need to be revised to reflect local antimicrobial susceptibility testing in line with recent findings and universal guidelines [22]

Geographical distribution of cases shows that, residents of the Cape Coast Metropolis and the Komenda-Edina-Eguafo-Abirem district were mostly affected by the outbreak with the highest attack rates that is, 285.2 and 23.4 per 100,00 population respectively. These communities were the most densely populated communities in comparison with all the other 5 affected districts. Studies have revealed that the risk of cholera is high amongst communities with slum settlements and densely populated communities, and it’s also high with communities in close proximity to dump sites and proximity to potentially contaminated surface water bodies and the spatial neighbors of communities [25].

The observed non-random distribution and sustained transmission of cholera is most likely influenced by demographic factors such as urbanization and overcrowding in the two districts. The risk of cholera infection is also high when majority of the people do not have access to good sanitation facilities; drink from rivers and wells; and when migration is high, this situation has been well documented in various studies conducted in the Central Region [18]. A significant association between the district of residence and the rapid diagnostic test results of cases (p-value=0.003), and that between the district of residence again and the final case classification (p-value=0.006), buttresses the fact that there was a certain pattern in the geographical distribution of cases, with most cases from the densely populated Cape Coast Metropolis and Komenda-Edina-Eguafo-Abirem testing positive for cholera and as well as culture confirmed to be cholera cases. As recommended by Badu Osei in 2010, in a study on the spatial statistics of epidemic data using a case study of cholera epidemiology in Ghana, it is necessary to prompt health officials and policy makers to execute measures to prevent fecal contamination of surface water bodies to prevent future cholera outbreaks in the Central Region. To achieve this; a house-to-house collection of solid waste should be extended to reduce the dependency on open space dumps; the Metropolitan Assembly should enforce the bylaws to prevent industrial pollution of surface water bodies. Also, the Metropolitan Assembly should increase the provision of good public sanitation facilities, such as flush toilets or water closets rather than the existing ventilated pit latrines. They should also draw and implement bylaws that will enforce landlords to provide toilet facilities for tenants in their houses; the Ghana Water Company Limited (GWCL) should improve the water distribution system so as to ensure constant (24 hours a day) supply of treated piped water to all inhabitants in the metropolis [25].The present study brings to the fore the challenges faced by developing countries in primarily poor socio-economic development, efficient town planning and creating effective surveillance as well as preparedness and response to cholera outbreaks. With respect to the nature of this outbreak and how diseases spread quickly, there is the need for an effective surveillance system with the capacity to timely and appropriately respond to and contain cholera outbreaks locally before they spread to neighboring areas.

## CONCLUSION

This study demonstrated a propagated type of outbreak driven by person-to-person transmission of infections which was worsened by episodes of common-source infections. An overall attack rate of 67 cases per 100,000 population recorded was one of the highest in terms of cholera outbreaks that have occurred in Ghana. Sex distribution was almost equal with majority of cases recorded in age groups 15-24 years. Residents of the densely populated Cape Coast Metropolis and the Komenda-Edina-Eguafo-Abirem district were mostly affected by the outbreak. It is recommended that district and sub-district surveillance systems be initiated to facilitate a quicker and more timely response in future outbreaks to prevent spread of cholera disease, current and future development projects must be geared towards provision of designated dumping sites and provision of more water treatment plants, provision of regular community education on cholera signs and symptoms and prevention and treatment measures even in periods outside epidemics and implementation of policies and laws governing sanitation at the district, sub-district and community levels.

## Data Availability

All data produced in the present study are available upon reasonable request to the authors

## ETHICAL CONSIDERATIONS

An ethical waiver and a subsequent approval for access to existing data on all cholera cases in the Central Region during the outbreak was obtained from the Disease Surveillance Department of the Ghana Health Service. All information obtained for the study (age, sex, date of onset of illness, district of residence, diagnostic test and culture results) was safeguarded within the database of the programme of the Disease Surveillance Department of the Ghana Health Service.

## ACKNOWLEDGEMENTS

The authors wish to thank the Central Regional Health Directorates staff and the District Health Management Team for their kind support in making this paper possible.

